# Left ventricular hypertrophy, brain atrophy and cognitive decline in type 2 diabetes mellitus: Diabetes & Dementia (D2) cohort study

**DOI:** 10.64898/2026.08.31.26361868

**Authors:** Amy Brodtmann, Sheila K Patel, Carolina Restrepo, Mohamed Salah Khlif, Emilio Werden, Rachel Ellis, Shahad Alsawaf, Elif Ekinci, Piyush M Srivastava, Jay Ramchand, Richard J MacIsaac, Leonid Churilov, Louise M Burrell

## Abstract

**BACKGROUND:** People with type 2 diabetes mellitus (T2DM) are at higher risk of cerebral small vessel disease and left ventricular hypertrophy (LVH), potentially contributing to cognitive decline and dementia. We aimed to describe brain volume and cognitive trajectories over 2 years in a cohort of people with T2DM and to determine whether LVH causes increased brain atrophy and cognitive decline.

**METHODS:** Diabetes and Dementia (D2) study is a multicentre observational cohort study in Melbourne, Australia. Participants aged >50 years were recruited via 2 hospital outpatient clinics, 3 private clinics, and study advertisements. Participants with pre-existing cognitive impairment, life-limiting medical illness, and severe chronic renal impairment were excluded. Participants attended study visits for brain MRI, transthoracic echocardiography (TTE), and cognitive testing at baseline and 2 years. The exposure was LVH determined on baseline TTE. Pre-specified outcomes were total brain volume (TBV) change and cognitive decline (z-score change≤-1 in any cognitive domain) over 2 years. Regression analyses examined associations between baseline variables and outcomes. A causal inference approach was utilized using inverse probability of treatment weighting to standardize for confounding covariates, excluding participants for non-positivity on age and baseline TBV.

**RESULTS:** Participants were recruited 20May2016 to 20March2020: 2378 screened, 702 eligible, 196 consented, 150 baseline and 123 2-year assessments with complete MRI, TTE, and cognitive data (17.4% attrition). At baseline, LVH was associated with female sex, older age, lower educational attainment, lower mood, hypertension, obesity, beta-blocker use, and smaller TBV. Participants with baseline cognitive impairment exhibited greater brain atrophy. Lower educational attainment, hypertension, and lower baseline cognitive scores were associated with cognitive decline. Causal inference analysis included 62 participants with no LVH (20(32%) women; mean [SD]=66.9[5.9] years), and 31 with LVH (17(55%) women, 67.4[5.4] years). LVH caused lower TBV change: standardized mean difference (95% CI) 6.3 (0.1, 12.5) cm^3^, *P* = .048. LVH had no effect on cognitive decline.

**CONCLUSIONS:** Brain atrophy and cognitive decline were associated with baseline cognitive impairment. LVH caused less brain atrophy and cognitive decline in people with T2DM. We conclude that guideline-directed LVH therapies such as beta-blockers have both cardioprotective (remodelling) and neuroprotective effects.

**TRIAL REGISTRATION:** ACTRN12616000546459 UTN: U1111-1181-6659

**Clinical perspective:** *What is new?:*

- Causal inference modelling was used to ask whether left ventricular hypertrophy (LVH) caused increased brain atrophy and cognitive decline over 2 years
- LVH reversed in 40% of participants
- Increased brain atrophy was associated with baseline cognitive impairment
- Lower educational attainment, baseline cognitive scores and hypertension were associated with cognitive decline
- LVH caused less brain atrophy with no effect on cognitive decline

*What are the clinical implications?:*

- LVH therapies, especially â-blockers, may have both cardioprotective (remodelling) and neuroprotective effects
- Cognitive impairment in people with T2DM should prompt aggressive risk factor management to prevent brain atrophy and cognitive decline

## Introduction

People with type 2 diabetes mellitus (T2DM) have increased risk of dementia^1^. Many factors may contribute to this strong association, including increased stroke incidence, cerebral small vessel disease (CSVD)^2^ and the cross-association of mid-life obesity and T2DM. Cerebral small vessel disease manifests as greater white matter hyperintensity (WMH) volume and lacunar infarcts^2^, also known risk factors for dementia. T2DM is associated with deposition of Alzheimer disease (AD) pathologies^3^. Insulin resistance and impaired glycemic control have been associated with increased risk of dementia, although the mechanisms remain controversial^4^. Efforts to treat vascular risk factors, especially hypertension^5^, have been proposed to reduce global dementia burden^6^. Whilst intensive glycemic control has not been associated with reduced risk of dementia^7^, there is now evidence that some drugs commonly used in the treatment of diabetes may be protective against cognitive decline^8^ and have been linked to lower incidence of dementia^9^.

People with T2DM often have hypertension, associated with left ventricular dysfunction and left ventricular hypertrophy (LVH)^10,11^. LVH is a known, independent predictor of adverse cardiovascular outcomes^12^, especially stroke and heart failure^13^. LVH has also been hypothesized as an independent risk factor for cognitive impairment^14^, but conflicting associations between LVH, brain atrophy, cognitive decline and dementia have been reported. This may be in part that, given the co-relationships between CSVD, diabetes and hypertension, prior studies have not been structured to examine direct relationships between LVH, cognition, and brain atrophy. A 2017 meta-analysis of population-based studies showed an increased risk of cognitive impairment among subjects with LVH, even after adjusting for hypertension or blood pressure levels^15^.

Researchers have used either ECG-defined or TTE-defined LVH criteria. Increased rates of incident probable dementia and mild cognitive impairment were reported in people with T2DM and malignant LVH^16^. LVH was not associated with worse baseline cognitive function but was associated with steeper decline in cognitive function in older subjects, independent of cardiovascular risk factors and co-morbidities^17^. LVH was associated with an increased risk of incident dementia but not with additional cognitive decline in the Atherosclerosis Risk in Communities study^18^. LV remodelling and diastolic dysfunction were associated with accelerated executive function, but not memory, decline^19^. This conflicts with the Second Manifestations of Arterial Disease-Magnetic Resonance (SMART-MR) study findings, where an increase in hypertensive target organ damage, particularly albuminuria and impaired renal function, was associated with greater decline in memory, not executive performance^20^.

Echocardiographic measures are regarded as more sensitive and accurate than ECG. In a cross-sectional study of 400 elderly people, an inverse trend across left ventricular mass increase (LVMI) quartiles and MMSE scores was reported (i.e., greater LV mass, worse cognitive score), and 2-fold increase in dementia^21^. MRI measures of LV structure and function were used in the Multi-Ethnic Study of Atherosclerosis in 4999 people free of cardiovascular disease at baseline, followed for incident dementia for around 12 years. Higher LVMI and LV mass-to-volume ratio (concentric remodelling) were independently associated with incident dementia and impaired cognitive function, but other measures of LV function were not^22^. Haring et al. followed 721 adults from the Strong Heart Study and CDCAI (Cerebrovascular Disease and Its Consequences in American Indians) Study with high prevalence of cardiovascular disease over 17 years, finding that those with higher LV mass had marginally lower hippocampal and cognitive scores, largely explicable by survivor bias^23^. None of these studies solely recruited people with T2DM, most used cognitive screens rather than more sensitive neuropsychological tests, and examined associations rather than causal inference.

In the Diabetes and Dementia (D2) study^24^, we performed a cohort study with two aims. We first aimed to describe demographic, cardiometabolic, brain imaging and cognitive features of people with T2DM comparing those with and without LVH, and to describe associations with brain atrophy (i.e., larger total brain volume (TBV) reduction) and cognitive decline at 2 years.

Given the prior association studies and the high correlation of hypertension, LVH, accelerated brain aging and cognitive impairment, our second aim was to investigate whether LVH would itself cause brain atrophy and cognitive decline. We chose to perform a causal inference analysis of the observational data, investigating an average exposure effect of LVH on participants with LVH. We hypothesized that, in people with T2DM and LVH, having LVH would lead to greater brain atrophy and more cognitive decline at 2 years.

## Methods

### Study design, participants and procedures

The Diabetes and Dementia study (D2) study was a cohort study with detailed cardiovascular, cognitive and brain imaging phenotyping of participants with T2DM. The study design and procedures are published^24^, and the statistical analysis plan is available on the Australian and New Zealand Clinical Trials registry (ACTRN12616000546459). We report according to the STROBE reporting guideline for cohort studies^25^.

The study was prospectively registered with the Australian and New Zealand Clinical Trials registry. Participants were recruited from 2 metropolitan university hospital diabetes outpatient (Austin Health, St Vincent’s Hospital Melbourne) and 3 private endocrinology clinics in Melbourne, Australia. Informed written consent was obtained from the participants before any data collection. Participants were free to withdraw at any time. Ethics approval was received from the Human Research Ethics Committee of Austin Health (24 March 2016, HREC/15/Austin/490). Recruitment commenced at Austin Health and private endocrinology rooms in 2016 and 27 March 2019 for St Vincent’s Hospital Melbourne.

Participants were included with a specialist-confirmed diagnosis of T2DM, age >50 years, and no prior history of neurological or psychiatric disease. Exclusion criteria were known prior cognitive impairment, significant medical co-morbidities precluding 2-year study participation, or CKD≥Stage 4/eGFR<30 mL/min/1.73m^2^, to reduce confounds, given the strong association with severe renal impairment and cognitive impairment^26,27^.

Participants attended 2 study visits, at baseline and at 2 years^24^. Study procedures at each study visit included brain MRI, functional, cognitive and mood assessments, and transthoracic echocardiogram (TTE). Participants were also fitted with 7-day physical activity monitors^24^, but these data were not examined here. Sex was reported as assigned at birth, and participants were given options for gender including non-binary. Weight and height were measured to the nearest 0.1kg/0.1cm during the clinical assessment using digital scales and a wall-mounted stadiometer. Years of education were binarized as ≤/>12 years. Obesity was defined as a body mass index (BMI)≥30kg/m^2^. Charlson Comorbidity Index (CCI) scores were used to summarize comorbidity^28^.

LV mass was calculated by LV cavity dimensions and wall thickness at end-diastole via the 2015 American Society of Echocardiography (ASE) recommended formula^29^. Left ventricular mass was indexed to height in metres^2.7^, categorized into LVH using ASE convention^29^. LVH was defined as>49 g/m^2.7^ for male sex and>45 g/m^2.7^ for female sex, binarized as present/absent based on this cut-off. All participants with TTE at both timepoints were remeasured by 2 blinded expert raters who were not part of the investigator team (see Acknowledgements and Supplementary Methods). There was substantial agreement between the two (κ=0.613, p< 0.001). The consensus opinion was to retain original allocations of LVH/no LVH as per published protocol^14,24^.

3 Tesla MRI scans were acquired on Siemens Skyra (2016-20) and Prisma scanners (2020-22) (64-channel head-neck coil) at the Melbourne Brain Centre, Florey Institute. Longitudinal FreeSurfer (v7.3.2) processing pipeline was used to segment 1mm isotropic T1-weighted images including motion correction, removal of non-brain tissue, automated Talairach transformation, subcortical white matter and deep gray matter segmentation, intensity normalization, gray matter-white matter boundary tessellation, and automated topology correction. T2-weighted images improved pial surface segmentation. All segmentations were visually inspected and corrected as required.

Cognitive and mood assessments were delivered primarily face-to-face, with video testing available during the SARS-CoV-19 pandemic as required. Tools and tests for cognitive assessments covered major cognitive domains, detailed in our protocol^24^. Age-appropriate normative values (mean, standard deviation) were used to create composite z-scores for each of 6 cognitive domains, including attention, visuospatial ability, executive function, language, memory, and processing speed^24^. Cognitive impairment was defined as a z-score≤-1.5 in any cognitive domain at baseline. Mood was assessed using the Generalized Anxiety Disorder (GAD-7)^30^ questionnaire and Patient Health Questionnaire (PHQ-9)^31^.

### Main Outcomes and Measures

The exposure variable was the presence of LVH at baseline measured via TTE. Prespecified outcomes were relative TBV change between baseline (t_1_) and 2 years (t_2_), defined as [TBV(t_2_)-TBV(t_1_)]/TBV(t_1_)], and cognitive decline, defined as a change of ≤-1 in the z-score of any cognitive domain from baseline to 2-year assessment.

### Statistical methods

Higher LV mass and concentric geometric changes have been demonstrated in 35-40% of patients with T2DM^32^. We have reported higher percentages in people with moderately severe T2DM^10^. We estimated the sample size to ensure sufficient participant numbers for both outcome measures in the cohort study. We based our effect size on published brain volume and cortical thickness estimates performed on healthy controls, people with CSVD and neurodegenerative disease^33–36^. Using alpha=0.05 (two-sided), power=0.8 and an estimated correlation between baseline and 2-year outcome values of 0.1, we estimated that 140 (i.e., 70 each in 2 groups) participants would be required to observe the effect of 0.4 or larger. We estimated a total recruitment number of 168 participants as we expected 20% attrition due to death or non-participation.

Analyses were conducted using SPSS version 30 (IBM SPSS Statistics, IBM Corp, USA) and Stata v18BE (StataCorp, College Station, TX, USA). The statistical analysis plan is presented in the Supplement, Appendix 1. Analyses were conducted separately for two aims. In aim 1, baseline characteristics (demographic, imaging, comorbidities, cognition) for the whole cohort were compared between the participants with and without LVH. Overall, continuous data were reported as means with standard deviations (SD); non-normally distributed data were expressed as medians with interquartile ranges (IQR, 25^th^,75^th^ percentiles), and categorical variables are shown as No. (%). Baseline characteristics were compared based on the presence or absence of LVH using an independent samples *t*-test for normally distributed continuous variables, the Mann-Whitney *U* test for non-normal data, and Pearson’s χ^2^ test for categorical variables.

Associations between baseline variables (univariate analyses) and TBV change and cognitive decline at 2 years were investigated using linear regression for TBV change and binary logistic regression for cognitive decline.

For aim 2, a causal inference analyses for the average LVH effect on participants with LVH were conducted, using LVH at baseline (yes vs. no) as the exposure for prespecified outcomes. Potential confounders for the causal relationship between the exposure and the outcomes were identified via an *a priori* formulated causal hypotheses using directed acyclic graphs (DAGs, Supplementary Figures 1 and 2). Inverse probability of treatment weighting (IPTW) using propensity scores for LVH was performed based on age, obesity, and TBV at baseline as confounders for the TBV outcome, and based on age, obesity, TBV, and education≤12 years for the cognitive decline outcome. The comparability of the LVH and non-LVH participants was assessed appropriately, and participants violating non-positivity assumptions were excluded from the causal analyses.

Two secondary IPTW analyses were performed. The first included hypertensive status as a confounder, as measured on participants’ 24-hour ambulatory blood pressure (binarized as yes vs. no). The presence of Stage 1 hypertension was determined as mean 24-hour ambulatory systolic blood pressure>130 mmHg or diastolic pressure>80 mmHg, following current AHA definitions^37^.

In addition, we repeated the original IPTW analyses for change in TBV and cognitive decline over 2 years including only participants who persistently classified as LVH or non-LVH at both baseline and follow-up assessments. See Supplementary Methods for details. To estimate the causal effect of the LVH exposure on the outcomes, we fitted an IPTW-weighted linear regression model for absolute change in TBV and a binary logistic regression model for decline in cognition using *teffects* command in Stata v18BE. Corresponding effects were reported as mean difference for linear regression and standardized risk difference for logistic regression with respective 95% confidence intervals based on robust standard error estimation. Two-tailed *p*-values<0.05 were considered significant.

### Blinding

Investigators were blinded to outcome of interest. One study staff member was unblinded to LVH status (SKP) and knew medical history details. Study staff fitting ambulatory BP monitors, and performing cognitive assessments (CR, RE) and imaging analyses (MSK) were blinded to LVH status. Unblinding only occurred after completing the statistical analysis plan, database lock, and transmission of data to our biostatistical team (LC).

## Results

Recruitment was curtailed by the SARS-CoV-19 pandemic. The date of last participant enrolment was set as 31 May 2020 to comply with local restrictions. Two-year follow-up visits were completed by July 2022. Figure 1 shows the flowchart of participant numbers for aims 1 and 2. All participants identified as cisgender.

**Figure 1:**
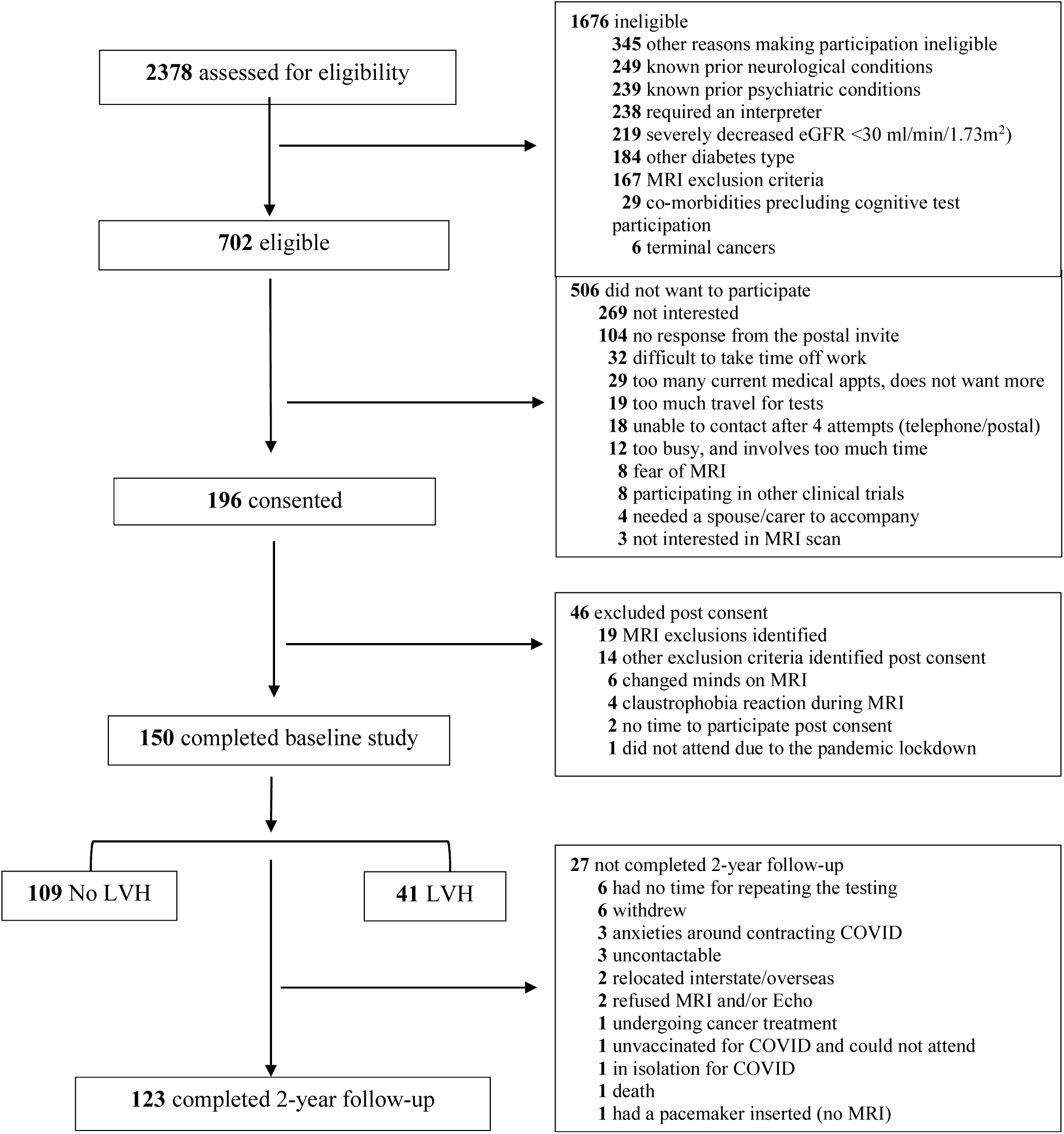
Study flow of eligibility, assessments and participants numbers.

Between 27 March 2016 and 31 May 2020, 2,378 participants were screened and 702(29.6%) were eligible. Of those eligible, 196(27.9%) consented and completed the pre-study screening and MRI safety check questionnaire, 46 patients were excluded or withdrew post-consent, 150 were enrolled and completed baseline assessments, 126 participants attended the 2-year study visit, with 123 completing all assessments. Only age and sex were collected for those who withdrew consent; there was no difference between those who withdrew (mean age 65.5(7.29) years, male 32(69.6%), female 14(30.4%)) compared to those who continued.

Baseline characteristics are summarized in Table 1 (Supplementary Table 1: ambulatory BP, echocardiographic, carotid intimal medial thickness, medications). There were 41 participants (27.3%) with LVH and 109 (72.7%) with no LVH. The overall mean participant age was 64.9[7] years, 64(43%) were female and 63(42.3%) participants had cognitive impairment at baseline. Participants with LVH were older (mean [SD] age, LVH 67.1[5.9], no LVH 64.1[7.3] years), had lower educational attainment (37% vs. 20%), higher BMI (LVH 33.1[6.3] vs. no LVH 29.4[5.1] kg/m^2^), and more likely to be female. Those with LVH had more comorbidities of hypertension and myocardial infarction, and more betablocker use (39% vs. 14%). Baseline TBV (1067.4[42.4] vs. 1085.3[61.5]cm^3^) and cognitive scores were lower in the LVH group with higher (worse) scores on depression screens (PHQ-9). More people with LVH had cognitive impairment at baseline (65.9% vs. 33.3%). HbA1_c_ and eGFR levels were not different between groups.

**Table 1:**
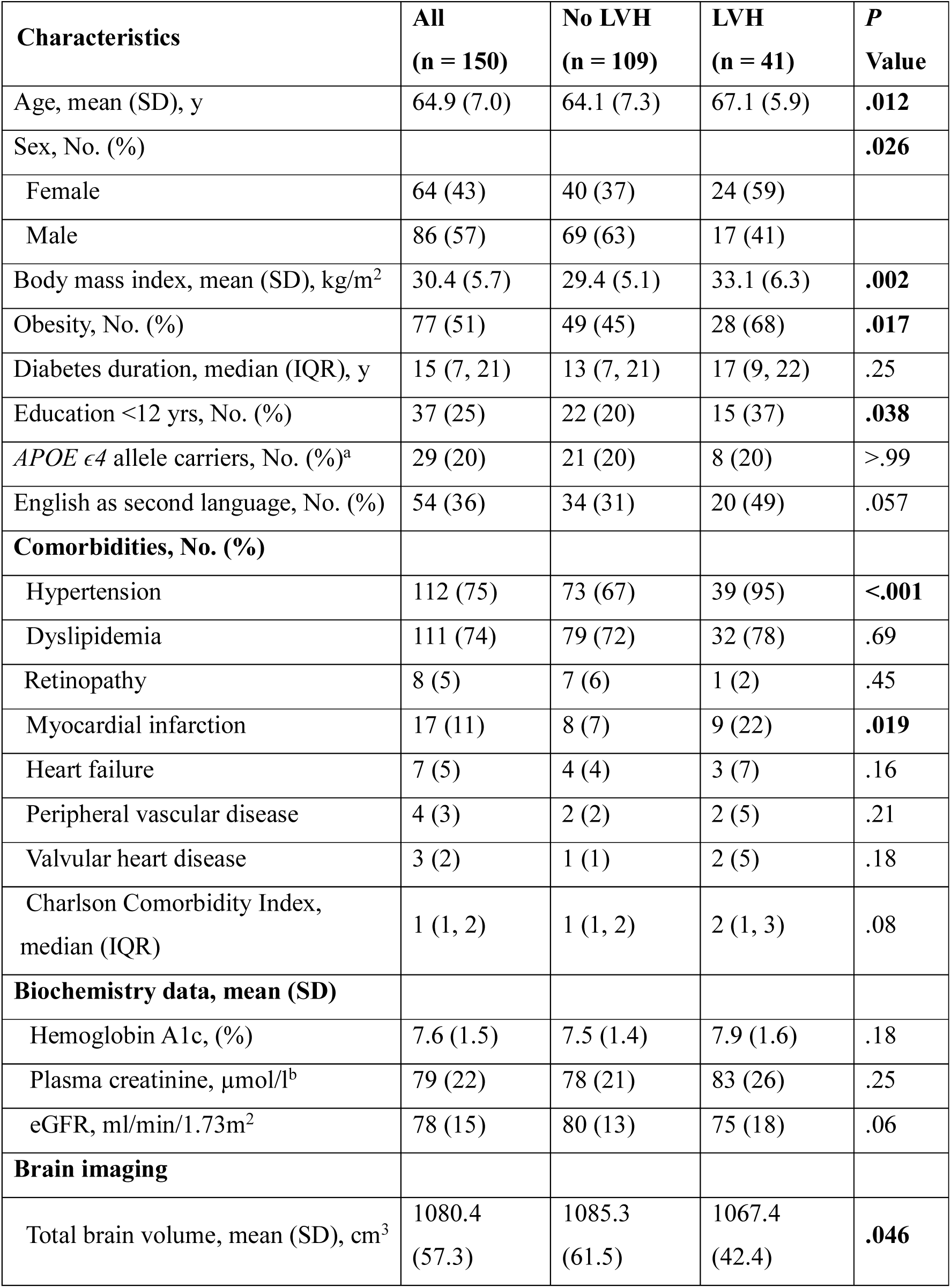
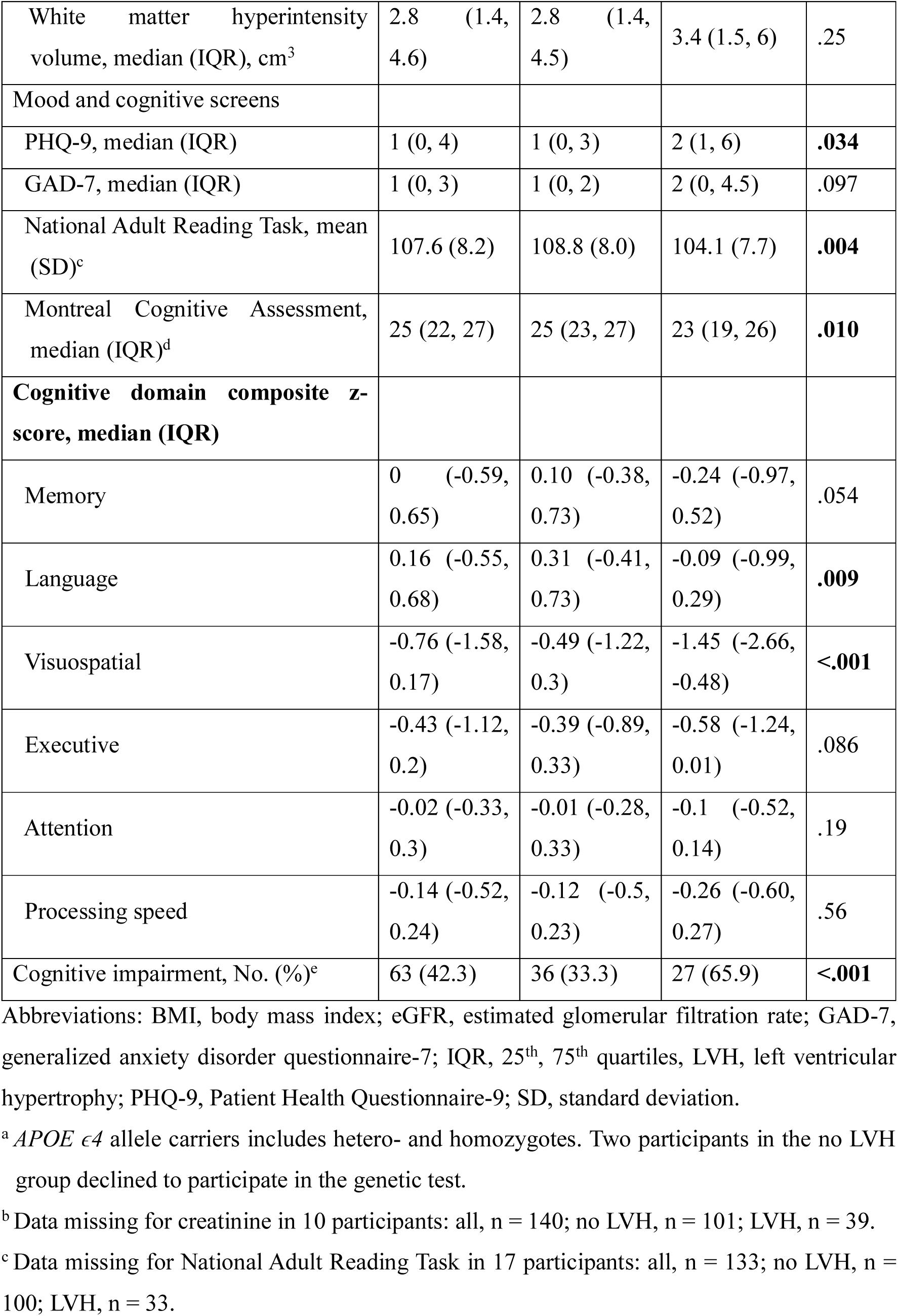
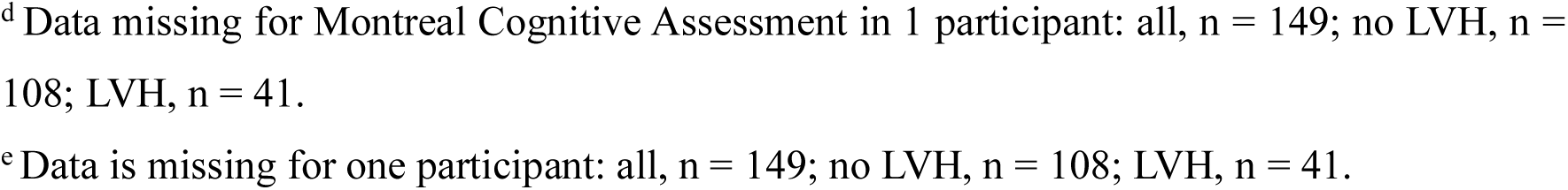
Baseline participant characteristics for the complete cohort, stratified by left ventricular hypertrophy status (Aim 1).

| Characteristics | All<br>(n = 150) | No LVH<br>(n = 109) | LVH<br>(n = 41) | P<br>Value |
| --- | --- | --- | --- | --- |
| Age, mean (SD), y | 64.9 (7.0) | 64.1 (7.3) | 67.1 (5.9) | **.012** |
| Sex, No. (%) |  |  |  | **.026** |
| Female | 64 (43) | 40 (37) | 24 (59) |  |
| Male | 86 (57) | 69 (63) | 17 (41) |  |
| Body mass index, mean (SD), kg/m<sup>2</sup> | 30.4 (5.7) | 29.4 (5.1) | 33.1 (6.3) | **.002** |
| Obesity, No. (%) | 77 (51) | 49 (45) | 28 (68) | **.017** |
| Diabetes duration, median (IQR), y | 15 (7, 21) | 13 (7, 21) | 17 (9, 22) | .25 |
| Education <12 yrs, No. (%) | 37 (25) | 22 (20) | 15 (37) | **.038** |
| *APOE* $\epsilon 4$ allele carriers, No. (%)<sup>a</sup> | 29 (20) | 21 (20) | 8 (20) | >.99 |
| English as second language, No. (%) | 54 (36) | 34 (31) | 20 (49) | .057 |
| **Comorbidities, No. (%)** |  |  |  |  |
| Hypertension | 112 (75) | 73 (67) | 39 (95) | **<.001** |
| Dyslipidemia | 111 (74) | 79 (72) | 32 (78) | .69 |
| Retinopathy | 8 (5) | 7 (6) | 1 (2) | .45 |
| Myocardial infarction | 17 (11) | 8 (7) | 9 (22) | **.019** |
| Heart failure | 7 (5) | 4 (4) | 3 (7) | .16 |
| Peripheral vascular disease | 4 (3) | 2 (2) | 2 (5) | .21 |
| Valvular heart disease | 3 (2) | 1 (1) | 2 (5) | .18 |
| Charlson Comorbidity Index,<br>median (IQR) | 1 (1, 2) | 1 (1, 2) | 2 (1, 3) | .08 |
| **Biochemistry data, mean (SD)** |  |  |  |  |
| Hemoglobin A1c, (%) | 7.6 (1.5) | 7.5 (1.4) | 7.9 (1.6) | .18 |
| Plasma creatinine, $\mu\text{mol/l}^b$ | 79 (22) | 78 (21) | 83 (26) | .25 |
| eGFR, ml/min/1.73m<sup>2</sup> | 78 (15) | 80 (13) | 75 (18) | .06 |
| **Brain imaging** |  |  |  |  |
| Total brain volume, mean (SD), cm<sup>3</sup> | 1080.4<br>(57.3) | 1085.3<br>(61.5) | 1067.4<br>(42.4) | **.046** |
| White matter hyperintensity volume, median (IQR), cm<sup>3</sup> | 2.8 (1.4, 4.6) | 2.8 (1.4, 4.5) | 3.4 (1.5, 6) | .25 |
| Mood and cognitive screens |  |  |  |  |
| PHQ-9, median (IQR) | 1 (0, 4) | 1 (0, 3) | 2 (1, 6) | **.034** |
| GAD-7, median (IQR) | 1 (0, 3) | 1 (0, 2) | 2 (0, 4.5) | .097 |
| National Adult Reading Task, mean (SD)<sup>c</sup> | 107.6 (8.2) | 108.8 (8.0) | 104.1 (7.7) | **.004** |
| Montreal Cognitive Assessment, median (IQR)<sup>d</sup> | 25 (22, 27) | 25 (23, 27) | 23 (19, 26) | **.010** |
| **Cognitive domain composite z-score, median (IQR)** |  |  |  |  |
| Memory | 0 (-0.59, 0.65) | 0.10 (-0.38, 0.73) | -0.24 (-0.97, 0.52) | .054 |
| Language | 0.16 (-0.55, 0.68) | 0.31 (-0.41, 0.73) | -0.09 (-0.99, 0.29) | **.009** |
| Visuospatial | -0.76 (-1.58, 0.17) | -0.49 (-1.22, 0.3) | -1.45 (-2.66, -0.48) | **<.001** |
| Executive | -0.43 (-1.12, 0.2) | -0.39 (-0.89, 0.33) | -0.58 (-1.24, 0.01) | .086 |
| Attention | -0.02 (-0.33, 0.3) | -0.01 (-0.28, 0.33) | -0.1 (-0.52, 0.14) | .19 |
| Processing speed | -0.14 (-0.52, 0.24) | -0.12 (-0.5, 0.23) | -0.26 (-0.60, 0.27) | .56 |
| Cognitive impairment, No. (%)<sup>e</sup> | 63 (42.3) | 36 (33.3) | 27 (65.9) | **<.001** |

Abbreviations: BMI, body mass index; eGFR, estimated glomerular filtration rate; GAD-7, generalized anxiety disorder questionnaire-7; IQR, 25<sup>th</sup>, 75<sup>th</sup> quartiles, LVH, left ventricular hypertrophy; PHQ-9, Patient Health Questionnaire-9; SD, standard deviation.
<sup>a</sup> *APOE* $\epsilon 4$ allele carriers includes hetero- and homozygotes. Two participants in the no LVH group declined to participate in the genetic test.
<sup>b</sup> Data missing for creatinine in 10 participants: all, n = 140; no LVH, n = 101; LVH, n = 39.
<sup>c</sup> Data missing for National Adult Reading Task in 17 participants: all, n = 133; no LVH, n = 100; LVH, n = 33.
<sup>d</sup> Data missing for Montreal Cognitive Assessment in 1 participant: all, n = 149; no LVH, n = 108; LVH, n = 41. <sup>e</sup> Data is missing for one participant: all, n = 149; no LVH, n = 108; LVH, n = 41.

One hundred twenty-six participants attended the 2-year study visit, 123 with complete data. This attrition rate of 18% was lower than the predicted rate of 20%^24^. No significant differences in age, sex, or educational status were observed between people who did or did not attend follow-up (Supplementary Table 2). Those who did not attend had more comorbidities of dyslipidemia and heart disease, more anticoagulant, diuretic, and betablocker use, and slower speed of processing at baseline.

LVH status of some changed over the 2 years: 5(5.4%) participants with no LVH at baseline developed LVH, and 12(40%) with LVH at baseline had no LVH at 2 years (reverse remodelling; Supplementary Table 3).

Age and sex were not associated with TBV change or cognitive decline (Table 2). TBV change over 2 years was greater (more brain atrophy) in participants with baseline cognitive impairment and with lower baseline TBV. Cognitive decline was associated with less educational attainment, more hypertension, and lower scores on the National Adult Reading Test (NART)^38^ and Montreal Cognitive Assessment (MoCA)^39^.

**Table 2:** Associations between selected demographic variables and prespecified outcomes (Aim 1). TBV change and cognitive decline over 2 years. Bold values denote significant differences.

|  | Total brain volume, cm<sup>3</sup> | Cognitive decline |
| --- | --- | --- |
| Characteristics | Rate of change<sup>a</sup> or<br>Mean difference<sup>b</sup> (95%<br>CI) | Odds ratio (95% CI) |
| Age (per 1 year) | -0.20 [-0.59 to 0.19] | 1.03 [0.98 to 1.08] |
| Sex |  |  |
| Female | NA | 1.0 [Reference] |
| Male | 0.37 [-5.15 to 5.89] | 0.98 [0.48 to 2.00] |
| Body mass index (per 1 kg/m<sup>2c</sup>) | 0.05 [-0.41 to 0.51] | 0.99 [0.93 to 1.05] |
| Diabetes duration (per 1 year) | 0.01 [-0.27 to 0.28] | 1.02 [0.98 to 1.05] |
| Education<sup>d</sup> |  |  |
| >12 years | NA | 1.0 [Reference] |
| <12 years | -3.88 [-10.31 to 2.55] | **2.57 [1.05 to 6.26]** |
| APOE ε4 allele carriers<sup>e</sup> |  |  |
| ε3 allele carriers | NA | 1.0 [Reference] |
| ε4 allele carriers | -0.12 [-7.11 to 6.87] | 1.13 [0.45 to 2.88] |
| English as second language |  |  |
| No | NA | 1.0 [Reference] |
| Yes | 1.63 [-4.04 to 7.31] | 1.70 [0.80 to 3.58] |
| Hypertension |  |  |
| No | NA | 1.0 [Reference] |
| Yes | 0.68 [-5.56 to 6.93] | **2.39 [1.02 to 5.59]** |
| Charlson Comorbidity Index, CCI<sup>#</sup> | -0.31 [-3.00 to 2.37] | 0.99 [0.71 to 1.39] |
| Brain imaging |  |  |
| Total brain volume, cm<sup>3</sup> | **-0.05 [-0.09 to -0.002]** | 1.00 [1.00 to 1.01] |
| White matter hyperintensity<br>volume, cm<sup>3</sup> | -0.21 [-0.73 to 0.31] | 0.96 [0.89 to 1.03] |
| Mood and cognitive screens |  |  |
| PHQ-9<sup>#</sup> | -0.08 [-0.86 to 0.70] | 1.02 [0.92 to 1.13] |
| GAD-7<sup>#</sup> | 0.66 [-0.32 to 1.63] | 1.06 [0.93 to 1.21] |
| National Adult Reading Task,<br>NART<sup>#f</sup> | -0.11 [-0.46 to 0.25] | **0.93 [0.88 to 0.98]** |
| Montreal Cognitive Assessment,<br>MoCA<sup>#</sup> | -0.10 [-0.88 to 0.68] | **0.83 [0.74 to 0.94]** |
| Baseline cognitive impairment<sup>g</sup> |  |  |
| No | NA | 1.0 [Reference] |
| Yes | **-6.96 [-12.34 to -1.58]** | 1.52 [0.74 to 3.13] |

Abbreviations: BMI, body mass index; GAD-7, generalized anxiety disorder questionnaire-7;
NA, not applicable; PHQ-9, Patient Health Questionnaire-9; # unit of change is per score unit
for these values, i.e., per 1 unit change increase for CCI, PHQ-9, GAD-7, NART, MoCA.
<sup>a</sup> Rate of change per unit increase for continuous variables.
<sup>b</sup> Mean difference for categorical variables.
<sup>c</sup> Body mass index calculated as weight in kg divided by height in m<sup>2</sup>.
<sup>d</sup> Education is binarized to < 12 years or >12 years.
<sup>e</sup> *APOE* $\epsilon 4$ binarized as $\epsilon 4$ vs. non $\epsilon 4$ . Two participants in the no-LVH group declined to undergo the genetic test.
<sup>f</sup> Data missing for National Adult Reading Task in 13 participants, n = 110.
<sup>g</sup> Presence of cognitive impairment was binarized to no/yes.

Thirty participants were excluded from causal inference modelling (Aim 2) due to non-positivity for age (n=16) and TBV (n=14). Table 3 shows a comparison of those included before and after IPTW. Table 4 presents the results of the IPTW causal inference analysis. At two years, the change in TBV was higher (greater brain atrophy) for the no LVH group compared to those with LVH: absolute change mean difference in TBV was 6.3 cm^3^ (95%CI: 0.1, 12.5), *P*=.048. No significant difference in standardized risk for cognitive decline at 2 years was observed (0.16 (95%CI: -0.06, 0.39)).

**Table 3:**
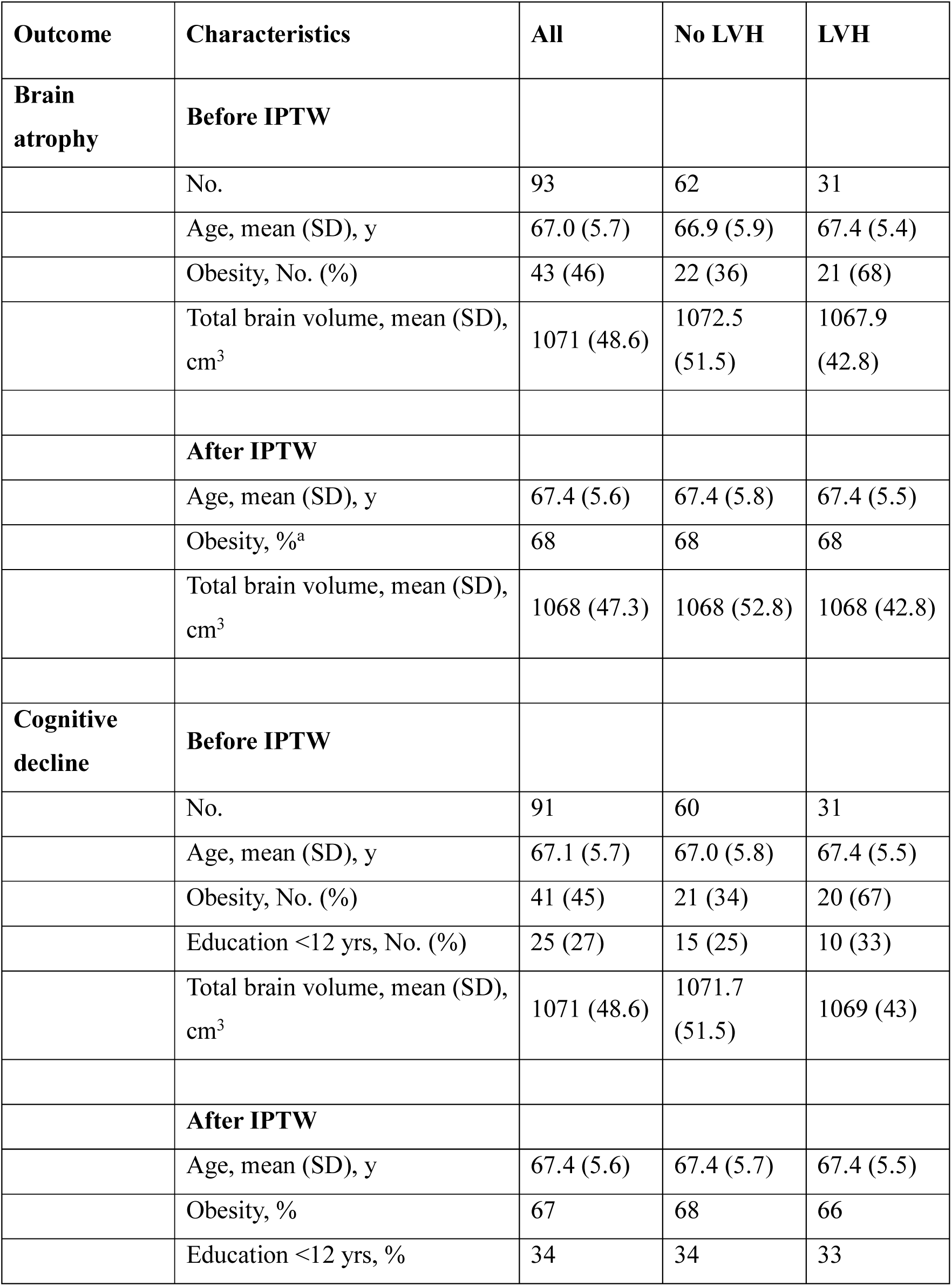
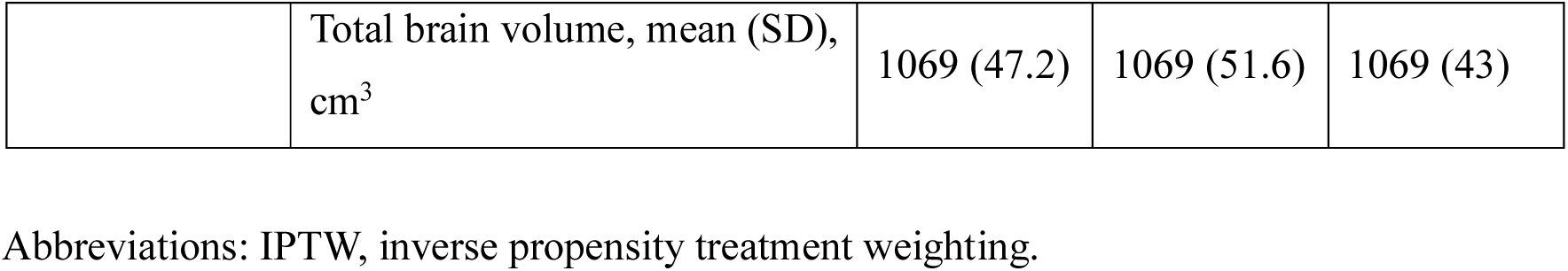
Baseline characteristics of participants included in causal inference analysis of the outcomes before and after inverse propensity treatment weighting (Aim 2).

**Table 4:**
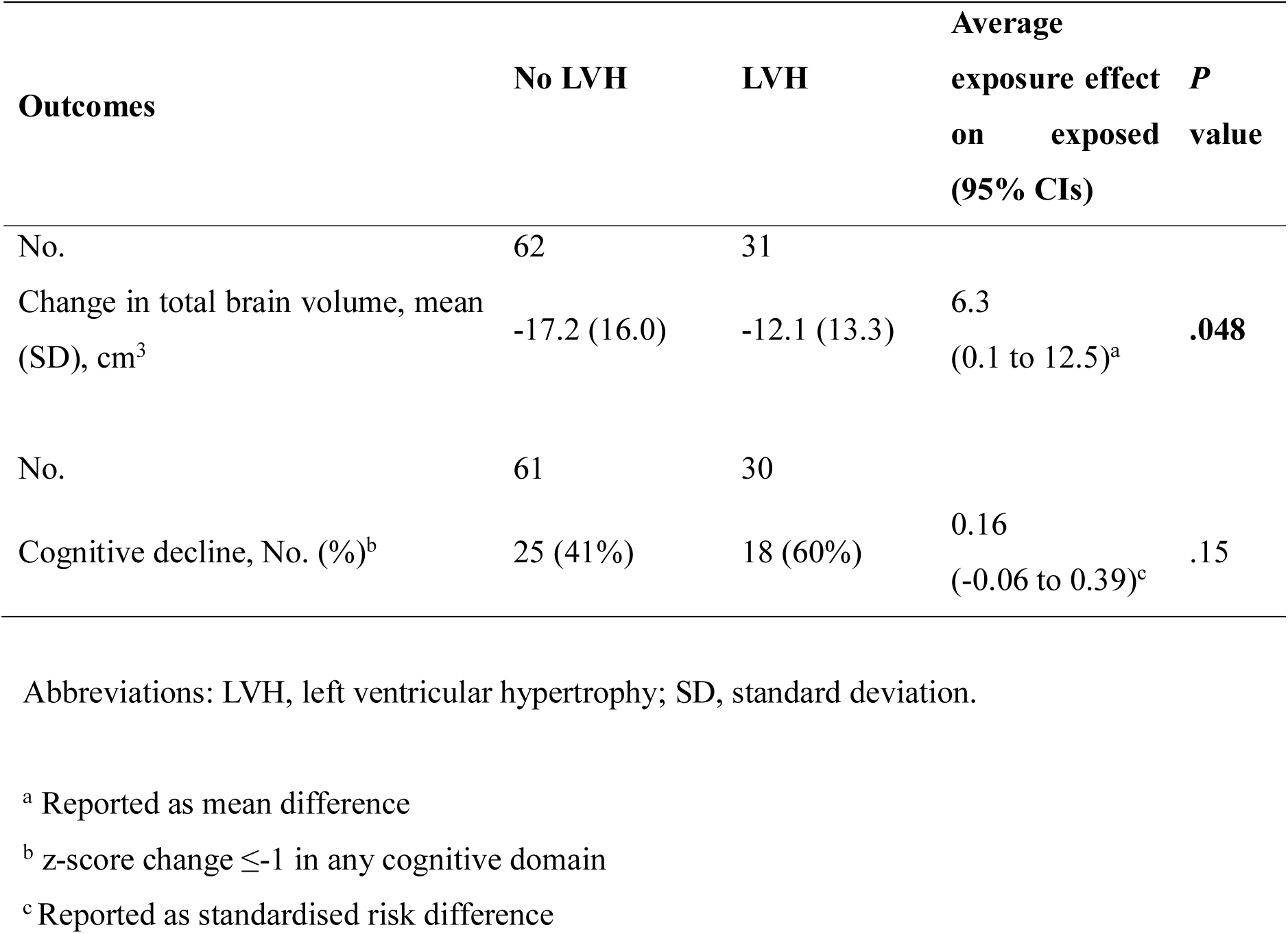
Causal inference analysis of outcomes at 2 years (Aim 2).

Results from our secondary analyses found that adding hypertension as a confounder did not alter the primary (aim 2) IPTW results. The reduction in TBV was still higher for the non-LVH (n=63) group (7.9 cm^3^ (95%CI: 1.1, 14.7), P=.024) compared to the LVH group (n=27). There was no significant difference in risk of cognitive decline at 2 years: 0.17 (95%CI: -0.08, 0.41), p=.19.

However, including only those participants with LVH/no LVH revealed no significant difference (4.4 cm^3^ (95%CI: -3.4, 12.1), p=.27) in change in TBV between participants with LVH (n=18) at both baseline and follow-up assessments and those who remained non-LVH (n=57). Difference in risk of cognitive decline again remained not significant: 0.07 (95%CI: - 0.27, 0.40), p=.69. Further details are provided in Supplementary Methods.

## Discussion

LVH is usually regarded as end organ damage: a long-term sequela of sustained hypertension, strongly associated with female sex, older age, and obesity^40^, as well as markers of cerebral small vessel disease^41^. In people with T2DM, we found that those with LVH were older, more likely to be women with lower educational attainment, more comorbidities, lower baseline cognitive scores and TBV, and higher depression scores. More people with LVH were classified as having cognitive impairment at baseline. That is, LVH correlated with the underlying drivers of both cardio- and cerebrovascular small vessel disease and dementia: hypertension, obesity, age, T2DM^42^.

Greater brain atrophy was associated with the presence of baseline cognitive impairment and lower TBV. We did not observe an association between older age and female sex with brain atrophy or cognitive decline. Cognitive decline was associated with lower educational attainment, lower baseline cognitive scores, and hypertension, all known risk factors for dementia. Our findings suggest that the drivers of brain atrophy have already manifested their effects at study enrolment in multiple end-organs, with LVH being one end-organ manifestation of these same risk factors.

In our causal inference modelling, we found that LVH caused less brain atrophy over 2 years, and there was no difference in cognitive decline between those with and without LVH. In our secondary analyses, these findings remained even after including current hypertension measured on 24-hour ambulatory BP monitoring as a confounder. Even when only those whose LVH status remained stable (i.e., LVH at both timepoints) were included in a further exploratory model, there was no difference between groups in either brain atrophy and cognitive decline. This was counter to prior findings from some population-based studies and to our *a priori* hypothesis.

Why would the exposure of LVH cause less brain atrophy? There are many potential explanations for this null hypothesis. The first is that our small sample size and relatively short follow-up of 2 years did not allow the detection of real effects. Further reducing the sample size via trimming for non-positivity may have also affected findings. Unlike most prior population studies, our sample only included people with T2DM. However, people with LVH differed in terms of baseline medication use, many of which are known to have both cardioprotective effects, via reverse remodelling, and neuroprotective effects^43^. Angiotensin receptor blockers (ARBs) and β-blockers, both recommended therapies for people with hypertension and LVH, have been associated with reduced dementia risk in older people^44^. β-blocker use is associated with reduced AD pathologies in elderly patients^45^. More people with LVH in our study were on β-blockers, but we did not find differences between groups in any other drug class. This may be because the use of β-blockers in people with LVH and coronary heart disease is recommended. More people with LVH in our study had a history of coronary heart disease or heart failure. β-blockers may confer neuroprotective benefits in a post-myocardial infarction rat model^37^. While a 2020 meta-analysis found no association between dementia risk and anti-hypertensive class^5^, there is some evidence from neuropathological data that angiotensin-II stimulating antihypertensive medications, defined as angiotensin receptor blockers, dihydropyridine calcium channel blockers and thiazides, reduce Alzheimer’s disease pathologies^38^. There are also encouraging data that both older (metformin^47^) and newer (glucagon-like peptide-1 receptor agonists^48^) agents used in the management of people with T2DM have positive effects on both LV diastolic function, LV mass, and cognition.

Forty percent of our participants with LVH at baseline were classified as no LVH at 2 years. This may in part have arisen from re-classification due to LV mass values close to cut-offs, but our blinded remeasurement demonstrated good agreement with original rating (see Supplementary Methods). Prior studies showing positive associations between echocardiographic LV metrics and cognition have largely modelled cross-sectional LV mass as a continuous variable or divided via high and low quartiles. We have planned analyses using LV mass indices as continuous variable, as well as LV geometrics and measures of diastolic function as cardiac exposures within causal frameworks^24^.

Our study has several limitations. Restrictions imposed during the COVID-19 pandemic were very strict in Melbourne, meaning that recruitment was curtailed. We had fewer people with LVH than expected based on prior national^10^ and international reports^49^, which may reflect better treatments for both hypertension and obesity. Whilst our participants had detailed phenotyping, not every biomarker was assessed, i.e., specific associations between high-sensitivity cardiac troponin or N-terminal pro-B-type natriuretic peptide, LVH and cognition were not examined. Our small sample size relative to other larger population studies and RCTs may also limit generalizability, especially as many participants were excluded due to MRI criteria. With only 91 participants and 43 events, the cognitive decline analyses are likely underpowered, given the wide confidence intervals (−0.06 to 0.39), and our follow-up interval of only 2 years is relatively short for a cognitive impairment study.

However, our study has several strengths. The use of a causal inference model and characterisation of LVH via TTE allowed assessment of whether LVH was driving cognitive decline and brain atrophy. Our participants were from a range of cultural and socioeconomic backgrounds, recruited both from public hospital clinics, where often people with more severe and chronic T2DM are seen, and from private endocrinology rooms. The close phenotyping, including high-resolution MRI, cardiac metrics, blood and cognitive testing at each timepoint allowed us to probe cognitive and structural imaging associations at baseline and to form a uniquely detailed cohort of people with T2DM. This will allow examination of other impacts on cardiac measures, cognition and brain structure in future studies.

## Conclusions

In our 2-year longitudinal study of people with T2DM, greater brain atrophy was associated with baseline cognitive impairment. Cognitive decline was associated with a lower level of education, lower baseline cognitive scores, and hypertension. LVH caused less brain atrophy and did not impact cognitive decline. Anti-hypertensive therapies, especially β-blockers, were used more in our LVH group and may have both cardioprotective (remodelling) and neuroprotective effects. Cognitive impairment in people with T2DM should prompt aggressive risk factor management to prevent brain atrophy and cognitive decline.

## Data Availability

Data collected for the study, including individual participant data and a data dictionary defining each field in the set, will be made available upon reasonable request. Proposals should be sent to the corresponding author for approval from the Steering Committee. Deidentified participant data, data dictionary, and imaging analysis pipelines could be shared from Jan 1, 2027. The study protocols, statistical analysis plan, and informed consent form are available from the Australian and New Zealand Clinical Trials website ACTRN12616000546459.

https://anzctr.org.au/Trial/Registration/TrialReview.aspx?ACTRN=ACTRN12616000546459

## Acknowledgements

We thank all our participants for their time. We would also like to thank the many contributors to our study: the D2 study co-ordination team, including Dr Venesha Rethnam, Rebecca Singleton, and Amanda Shanks; Diabetes Outpatient clinic staff at Heidelberg Repatriation Hospital and St Vincent’s Hospital Melbourne; imaging staff at the Melbourne Brain Centre; the cardiology department at the Austin Hospital; Advara Heartcare, Warringal Private Hospital for conducting echocardiograms and electrocardiograms; vascular sonographers Joanne Leech and Joel Ashworth at the Neurodiagnostic Laboratory, Austin Hospital. Thanks to Professor Christopher O’Callaghan for his guidance and training in fitting and conducting ambulatory blood pressure measurements. A special thanks to cardiology fellow Dr Arpudh Anandaraj and cardiac technologist Simon Undrill for conducting the double-blinded rating of echocardiograms.

## References

1. Biessels GJ, Despa F. Cognitive decline and dementia in diabetes mellitus: mechanisms and clinical implications. Nature reviews Endocrinology. 2018;14:591–604. doi: 10.1038/s41574-018-0048-7

2. Duering M, Biessels GJ, Brodtmann A, Chen C, Cordonnier C, de Leeuw FE, Debette S, Frayne R, Jouvent E, Rost NS, et al. Neuroimaging standards for research into small vessel disease-advances since 2013. Lancet Neurol. 2023;22:602–618. doi: 10.1016/S1474-4422(23)00131-X

3. van Gils V, Rizzo M, Cote J, Viechtbauer W, Fanelli G, Salas-Salvado J, Wimberley T, Bullo M, Fernandez-Aranda F, Dalsgaard S, et al. The association of glucose metabolism measures and diabetes status with Alzheimer’s disease biomarkers of amyloid and tau: A systematic review and meta-analysis. Neurosci Biobehav Rev. 2024;159:105604. doi: 10.1016/j.neubiorev.2024.105604

4. Lemche E, Killick R, Mitchell J, Caton PW, Choudhary P, Howard JK. Molecular mechanisms linking type 2 diabetes mellitus and late-onset Alzheimer’s disease: A systematic review and qualitative meta-analysis. Neurobiol Dis. 2024;196:106485. doi: 10.1016/j.nbd.2024.106485

5. Ding J, Davis-Plourde KL, Sedaghat S, Tully PJ, Wang W, Phillips C, Pase MP, Himali JJ, Gwen Windham B, Griswold M, et al. Antihypertensive medications and risk for incident dementia and Alzheimer’s disease: a meta-analysis of individual participant data from prospective cohort studies. Lancet Neurol. 2020;19:61–70. doi: 10.1016/S1474-4422(19)30393-X

6. Gladman JT, Corriveau RA, Debette S, Dichgans M, Greenberg SM, Sachdev PS, Wardlaw JM, Biessels GJ. Vascular contributions to cognitive impairment and dementia: Research consortia that focus on etiology and treatable targets to lessen the burden of dementia worldwide. Alzheimers Dement (N Y). 2019;5:789–796. doi: 10.1016/j.trci.2019.09.017

7. Areosa Sastre A, Vernooij RW, Gonzalez-Colaco Harmand M, Martinez G. Effect of the treatment of Type 2 diabetes mellitus on the development of cognitive impairment and dementia. The Cochrane database of systematic reviews. 2017;6:CD003804. doi: 10.1002/14651858.CD003804.pub2

8. Kuate Defo A, Bakula V, Pisaturo A, Labos C, Wing SS, Daskalopoulou SS. Diabetes, antidiabetic medications and risk of dementia: A systematic umbrella review and meta-analysis. Diabetes Obes Metab. 2024;26:441–462. doi: 10.1111/dom.15331

9. Milori PH, Armelin LM, Mutarelli A, Caramelli P. Incidence of dementia in patients with type 2 diabetes using SGLT2 inhibitors versus GLP-1 receptor agonists or DPP-4 inhibitors: A systematic review and meta-analysis of cohort studies. J Alzheimers Dis. 2025:13872877251395086. doi: 10.1177/13872877251395086

10. Srivastava PM, Calafiore P, Macisaac RJ, Patel SK, Thomas MC, Jerums G, Burrell LM. Prevalence and predictors of cardiac hypertrophy and dysfunction in patients with Type 2 diabetes. Clin Sci (Lond). 2008;114:313–320. doi: 10.1042/CS20070261

11. Patel SK, Wai B, Lang CC, Levin D, Palmer CNA, Parry HM, Velkoska E, Harrap SB, Srivastava PM, Burrell LM. Genetic Variation in Kruppel like Factor 15 Is Associated with Left Ventricular Hypertrophy in Patients with Type 2 Diabetes: Discovery and Replication Cohorts. EBioMedicine. 2017;18:171–178. doi: 10.1016/j.ebiom.2017.03.036

12. Laurent S, Agabiti-Rosei C, Bruno RM, Rizzoni D. Microcirculation and Macrocirculation in Hypertension: A Dangerous Cross-Link? Hypertension. 2022;79:479–490. doi: 10.1161/HYPERTENSIONAHA.121.17962

13. Lieb W, Xanthakis V, Sullivan LM, Aragam J, Pencina MJ, Larson MG, Benjamin EJ, Vasan RS. Longitudinal tracking of left ventricular mass over the adult life course: clinical correlates of short- and long-term change in the framingham offspring study. Circulation. 2009;119:3085–3092. doi: 10.1161/CIRCULATIONAHA.108.824243

14. Restrepo C, Patel SK, Rethnam V, Werden E, Ramchand J, Churilov L, Burrell LM, Brodtmann A. Left ventricular hypertrophy and cognitive function: a systematic review. J Hum Hypertens. 2018;32:171–179. doi: 10.1038/s41371-017-0023-0

15. Georgakis MK, Synetos A, Mihas C, Karalexi MA, Tousoulis D, Seshadri S, Petridou ET. Left ventricular hypertrophy in association with cognitive impairment: a systematic review and meta-analysis. Hypertens Res. 2017;40:696–709. doi: 10.1038/hr.2017.11

16. Kazibwe R, Ahmad MI, Hughes TM, Chen LY, Soliman EZ. Malignant left ventricular hypertrophy and risk of cognitive impairment in SPRINT MIND trial. Am Heart J. 2024;276:31–38. doi: 10.1016/j.ahj.2024.07.012

17. Mahinrad S, Vriend AE, Jukema JW, van Heemst D, Sattar N, Blauw GJ, Macfarlane PW, Clark EN, de Craen AJM, Sabayan B. Left Ventricular Hypertrophy and Cognitive Decline in Old Age. J Alzheimers Dis. 2017;58:275–283. doi: 10.3233/JAD-161150

18. Norby FL, Chen LY, Soliman EZ, Gottesman RF, Mosley TH, Alonso A. Association of left ventricular hypertrophy with cognitive decline and dementia risk over 20 years: The Atherosclerosis Risk In Communities-Neurocognitive Study (ARIC-NCS). Am Heart J. 2018;204:58–67. doi: 10.1016/j.ahj.2018.07.007

19. Dehghan A, Giugni FR, Bartholdy KV, Yang Y, Lamberson V, Wang W, Claggett B, Hajjar I, Mosley T, Palta P, et al. Left ventricular remodeling and diastolic dysfunction predict cognitive decline in older adults. Alzheimers Dement. 2026;22:e71089. doi: 10.1002/alz.71089

20. van der Veen PH, Geerlings MI, Visseren FL, Nathoe HM, Mali WP, van der Graaf Y, Muller M, Group SS. Hypertensive Target Organ Damage and Longitudinal Changes in Brain Structure and Function: The Second Manifestations of Arterial Disease-Magnetic Resonance Study. Hypertension. 2015;66:1152–1158. doi: 10.1161/HYPERTENSIONAHA.115.06268

21. Scuteri A, Coluccia R, Castello L, Nevola E, Brancati AM, Volpe M. Left ventricular mass increase is associated with cognitive decline and dementia in the elderly independently of blood pressure. Eur Heart J. 2009;30:1525–1529. doi: 10.1093/eurheartj/ehp133

22. Moazzami K, Ostovaneh MR, Ambale Venkatesh B, Habibi M, Yoneyama K, Wu C, Liu K, Pimenta I, Fitzpatrick A, Shea S, et al. Left Ventricular Hypertrophy and Remodeling and Risk of Cognitive Impairment and Dementia: MESA (Multi-Ethnic Study of Atherosclerosis). Hypertension. 2018;71:429–436. doi: 10.1161/HYPERTENSIONAHA.117.10289

23. Haring B, Omidpanah A, Suchy-Dicey AM, Best LG, Verney SP, Shibata DK, Cole SA, Ali T, Howard BV, Buchwald D, et al. Left Ventricular Mass, Brain Magnetic Resonance Imaging, and Cognitive Performance: Results From the Strong Heart Study. Hypertension. 2017;70:964–971. doi: 10.1161/HYPERTENSIONAHA.117.09807

24. Patel SK, Restrepo C, Werden E, Churilov L, Ekinci EI, Srivastava PM, Ramchand J, Wai B, Chambers B, O’Callaghan CJ, et al. Does left ventricular hypertrophy affect cognition and brain structural integrity in type 2 diabetes? Study design and rationale of the Diabetes and Dementia (D2) study. BMC Endocr Disord. 2017;17:24. doi: 10.1186/s12902-017-0173-7

25. von Elm E, Altman DG, Egger M, Pocock SJ, Gotzsche PC, Vandenbroucke JP, Initiative S. The Strengthening the Reporting of Observational Studies in Epidemiology (STROBE) statement: guidelines for reporting observational studies. Lancet. 2007;370:1453–1457. doi: 10.1016/S0140-6736(07)61602-X

26. Babroudi S, Tighiouart H, Schrauben SJ, Cohen JB, Fischer MJ, Rahman M, Hsu CY, Sozio SM, Weir M, Sarnak M, et al. Blood Pressure, Incident Cognitive Impairment, and Severity of CKD: Findings From the Chronic Renal Insufficiency Cohort (CRIC) Study. American journal of kidney diseases : the official journal of the National Kidney Foundation. 2023;82:443–453 e441. doi: 10.1053/j.ajkd.2023.03.012

27. Kalirao P, Pederson S, Foley RN, Kolste A, Tupper D, Zaun D, Buot V, Murray AM. Cognitive impairment in peritoneal dialysis patients. American journal of kidney diseases : the official journal of the National Kidney Foundation. 2011;57:612–620. doi: 10.1053/j.ajkd.2010.11.026

28. D’Hoore W, Sicotte C, Tilquin C. Risk adjustment in outcome assessment: the Charlson comorbidity index. Methods Inf Med. 1993;32:382–387.

29. Lang RM, Badano LP, Mor-Avi V, Afilalo J, Armstrong A, Ernande L, Flachskampf FA, Foster E, Goldstein SA, Kuznetsova T, et al. Recommendations for cardiac chamber quantification by echocardiography in adults: an update from the American Society of Echocardiography and the European Association of Cardiovascular Imaging. Journal of the American Society of Echocardiography : official publication of the American Society of Echocardiography. 2015;28:1–39 e14. doi: 10.1016/j.echo.2014.10.003

30. Spitzer RL, Kroenke K, Williams JB, Lowe B. A brief measure for assessing generalized anxiety disorder: the GAD-7. Archives of internal medicine. 2006;166:1092–1097. doi: 10.1001/archinte.166.10.1092

31. Kroenke K, Spitzer RL, Williams JB. The PHQ-9: validity of a brief depression severity measure. J Gen Intern Med. 2001;16:606–613.

32. Zanchetti A. Hypertension: Cardiac hypertrophy as a target of antihypertensive therapy. Nature reviews Cardiology. 2010;7:66–67. doi: 10.1038/nrcardio.2009.229

33. Chen K, Langbaum JB, Fleisher AS, Ayutyanont N, Reschke C, Lee W, Liu X, Bandy D, Alexander GE, Thompson PM, et al. Twelve-month metabolic declines in probable Alzheimer’s disease and amnestic mild cognitive impairment assessed using an empirically pre-defined statistical region-of-interest: findings from the Alzheimer’s Disease Neuroimaging Initiative. Neuroimage. 2010;51:654–664. doi: S1053-8119(10)00251-X [pii] 10.1016/j.neuroimage.2010.02.064

34. Hua X, Lee S, Hibar DP, Yanovsky I, Leow AD, Toga AW, Jack CR, Jr., Bernstein MA, Reiman EM, Harvey DJ, et al. Mapping Alzheimer’s disease progression in 1309 MRI scans: power estimates for different inter-scan intervals. Neuroimage. 2010;51:63–75. doi: S1053-8119(10)00143-6 [pii] 10.1016/j.neuroimage.2010.01.104

35. Leow AD, Yanovsky I, Parikshak N, Hua X, Lee S, Toga AW, Jack CR, Jr., Bernstein MA, Britson PJ, Gunter JL, et al. Alzheimer’s disease neuroimaging initiative: a one-year follow up study using tensor-based morphometry correlating degenerative rates, biomarkers and cognition. Neuroimage. 2009;45:645–655.

36. Brodtmann A, Puce A, Darby D, Donnan G. Regional fMRI brain activation does correlate with global brain volume. Brain research. 2009;1259:17–25. doi: 10.1016/j.brainres.2008.12.044

37. Writing Committee M, Jones DW, Ferdinand KC, Taler SJ, Johnson HM, Shimbo D, Abdalla M, Altieri MM, Bansal N, Bello NA, et al. 2025 AHA/ACC/AANP/AAPA/ABC/ACCP/ACPM/AGS/AMA/ASPC/NMA/PCNA/SGI M Guideline for the Prevention, Detection, Evaluation and Management of High Blood Pressure in Adults: A Report of the American College of Cardiology/American Heart Association Joint Committee on Clinical Practice Guidelines. Circulation. 2025;152:e114–e218. doi: 10.1161/CIR.0000000000001356

38. Nelson HE. The National Adult Reading Test (NART): test manual. In: NFER-Nelson; 1982.

39. Nasreddine ZS, Phillips NA, Bedirian V, Charbonneau S, Whitehead V, Collin I, Cummings JL, Chertkow H. The Montreal Cognitive Assessment, MoCA: a brief screening tool for mild cognitive impairment. J Am Geriatr Soc. 2005;53:695–699. doi: 10.1111/j.1532-5415.2005.53221.x

40. Cuspidi C, Rescaldani M, Sala C, Grassi G. Left-ventricular hypertrophy and obesity: a systematic review and meta-analysis of echocardiographic studies. J Hypertens. 2014;32:16–25. doi: 10.1097/HJH.0b013e328364fb58

41. Papadopoulos A, Palaiopanos K, Protogerou AP, Paraskevas GP, Tsivgoulis G, Georgakis MK. Left Ventricular Hypertrophy and Cerebral Small Vessel Disease: A Systematic Review and Meta-Analysis. Journal of stroke. 2020;22:206–224. doi: 10.5853/jos.2019.03335

42. Livingston G, Huntley J, Liu KY, Costafreda SG, Selbaek G, Alladi S, Ames D, Banerjee S, Burns A, Brayne C, et al. Dementia prevention, intervention, and care: 2024 report of the Lancet standing Commission. The Lancet 2024. doi: 10.1016/S0140-6736(24)01296-0

43. Mercanoglu G, Bamac OE, Sennazli G, Kalayci R, Mercanoglu F. Beyond the Heart: The Neuroprotective Potential of Nebivolol in Acute Myocardial Infarction. Life (Basel). 2025;15. doi: 10.3390/life15121880

44. Orchard SG, Zhou Z, Fravel M, Ryan J, Woods RL, Wolfe R, Shah RC, Murray A, Sood A, Reid CM, et al. Antihypertensive medications and dementia in older adults with hypertension. medRxiv. 2024. doi: 10.1101/2024.08.28.24312754

45. Lee S, Rajmohan R, Al-Darsani Z, Paganini-Hill A, Montine TJ, Corrada MM, Kawas C. Medical Histories Associated With Absence of Alzheimer Disease Neuropathologic Changes in the Oldest-Old: The 90+ Study. Neurology. 2025;104:e210107. doi: 10.1212/WNL.0000000000210107

46. Gray SL, Yu O, Gatto NM, Marcum ZA, Latimer CS, Postupna N, Su YR, Barthold D, van Dalen JW, Richard E, et al. Angiotensin II-Stimulating Antihypertensive Medications and Dementia-Related Neuropathology. JAMA Netw Open. 2026;9:e2559113. doi: 10.1001/jamanetworkopen.2025.59113

47. Azarboo A, Jalali S, Shaker F, Khalaji A, Assempoor R, Javidan A, Mohammadi Y, Bahrami O, Farahmandniya N, Cannavo A. The impact of metformin on myocardial hypertrophy: an updated systematic review, meta-analysis, and meta-regression of randomized controlled trials. Diabetol Metab Syndr. 2025. doi: 10.1186/s13098-025-02047-2

48. Zhang Y, Yang D, Jia Q, Yan J, An F. The effect of glucagon-like peptide-1 receptor agonists on cardiac function and structure in patients with or without type 2 diabetes mellitus: An updated systematic review and meta-analysis. Diabetes Obes Metab. 2024;26:2401–2411. doi: 10.1111/dom.15557

49. Lv J, Liu Y, Yan Y, Sun D, Fan L, Guo Y, Fernandez C, Bazzano L, He J, Li S, et al. Relationship Between Left Ventricular Hypertrophy and Diabetes Is Likely Bidirectional: A Temporality Analysis. J Am Heart Assoc. 2023;12:e028219. doi: 10.1161/JAHA.122.028219

